# Identifying High and Low Performing Emergency General Surgery Hospitals Using Direct Standardization

**DOI:** 10.1101/2024.02.23.24303292

**Authors:** Drew Goldberg, Luke Keele, Chris Wirtalla, Solomiya Syvyk, Rachel R. Kelz

## Abstract

**Importance:** Variation in outcomes for emergency general surgery conditions has been shown at the hospital level. Few have examined difference across hospitals for older adults who often present with the greatest risk. To date, no one has examined differences in the outcome for those undergoing operative and nonoperative treatment.

**Objective:** Identify high and low performing emergency general surgery (EGS) hospitals with risk-standardization to determine clinical performance differences as well as correlation between patients treated operatively and non-operatively.

**Design:** A retrospective cohort study with 30-day outcomes.

**Setting:** Nationwide study of acute care hospitals.

**Participants:** Medicare beneficiaries > 65.5 years old hospitalized for an emergency general surgery condition admitted from July 1, 2015 to June 30, 2018.

**Exposure:** Unique hospital identification.

**Main outcome:** A composite metric of 30-day mortality, adverse events, prolonged length of stay, and readmission.

**Results:** There were 536,284 total patients with a mean age of 74.4 ± 12.2 years, 55% female, 84% white with average claims-based frailty index of 0.16 ± 0.06 and mean comorbidity count of 3.57 ± 2.46. Amongst the 1866 hospitals identified, there were 3 best performing and 11 worst performing hospitals. There were weak correlations between operative and non-operative for mortality (0.10), adverse events rates (0.21), prolonged length of stay (0.32), and readmissions (0.18) at the hospital level (all p<0.001).

**Conclusions and Relevance:** Significant variation exists in EGS hospital performance with best ranked hospitals out-performing worst ranked hospitals on adverse event, mortality, prolonged length of stay and readmission. There is little association between patient outcomes for those treated with operative and non-operative care.

## Introduction

Emergency general surgery (EGS) hospitalizations comprise a significant proportion of total admissions in the United States with both operative and non-operative treatment options available to EGS patients.^1^ Given variation across both patient-level and hospital-level risk means, certain patients may do better being directed to particular hospitals.^2^ As such, measuring quality amongst EGS hospitals can provide insights into where patients would have the best outcome.

One method to capture variability across EGS hospitals performance includes ranking hospitals according to both mortality and non-mortality measures.^3^ Given the heterogeneity in both patient and hospital characteristics of EGS-treated populations, such an analysis provides meaningful insight into consistencies or deviations in both treatment pathways and resource utilization.

We sought to use a novel patient risk-adjustment method to rank EGS hospitals according to mortality, readmission, length of stay, and adverse events. We looked to identify both high and low performers amongst hospitals to understand unique attributes of those hospitals to serve as signals for hospital selection. We then sought to determine if patient outcomes were correlated among operative and non-operative patient populations.

## Methods

Patients with a principal diagnosis of an EGS conditions who were Medicare (fee-for-service) beneficiaries 65.5 years of age and older, and admitted through the emergency department to a non-federal, acute care hospital between July 1, 2015 and June 30, 2018 were included. EGS conditions were identified using International Classification of Disease, Ninth or Tenth Revision, Clinical Modification codes.^4^ Patient comorbidities were classified and analyzed according to previously published methods.^4–6^ The main outcome was a composite of 30-day adverse events defined by death, prolonged length of stay or readmission within 30-days of hospital discharge.^7^ Death, prolonged length of stay and readmission within 30-days were examined as secondary outcomes. Hospital characteristics were extracted from the American Hospital Association and mean star rating from the Medicare Hospital Compare tool.^8,9^

To control for differences in patient populations across hospitals that could account for estimated differences in hospital outcomes, we performed direct standardization using balancing weights on 71 indicators. Specifically, we standardize each hospitals’ patient population to the overall patient population.^10^ Balancing weights are a computational generalization of inverse propensity score weight that we use to create hospitals with patient populations with similar covariate distributions. Balancing weights assign a scalar weight to each patient such that we can take hospital level weighted averages that account for differences across hospital populations. We then generated hospital level rankings using an Empirical Bayes estimator that corrects for variation in the size of each hospital’s patient population. Prior research has shown that a balancing weights approach to direct standardization reduces bias due to observed differences between the comparison in patient populations which increasing precision by reducing the loss of sample size.^10^ We also identified the set of hospitals with a high probability of having a high or low rank relative to the overall distribution. Standard and rank correlations of hospitals were examined between performance for operative and nonoperative treatments. All statistical analyses were conducted in R version 4.1.1.

## Results

There were 536,284 total patients with a mean age of 74.4 ± 12.2 years and frailty index of 0.16 ± 0.06. Most patients were female (55%), white (84%), and had 3+ comorbid conditions (61%). Ten percent of the population was Black and 2% Hispanic.

Among the 1866 hospitals identified, 3 best performing hospitals were identified for all patients. The mean hospital compare star rating was 3.7 ± 0.9. There were 11 identified worst performing hospitals with a mean hospital star rating of 2.4 ± 1.1. For operative treatment, there were no distinctly best performing hospitals and 3 worst performing hospitals identified. Of these 3 hospitals, the average hospital stars rating was 2.9 ± 0.8. For non-operative treatment, there were 6 identified best performing and 2 worst performing hospitals. The best performing non-operative hospitals had a mean star rating of 2.8 ± 0.5. Amongst the hospitals that were worst performing at non-operative treatment (n=2), there was an average star rating of 3.0 ± 1.4. There were no apparent differences in hospital characteristics across hospitals by performance. (Table 1)

**Table 1:**
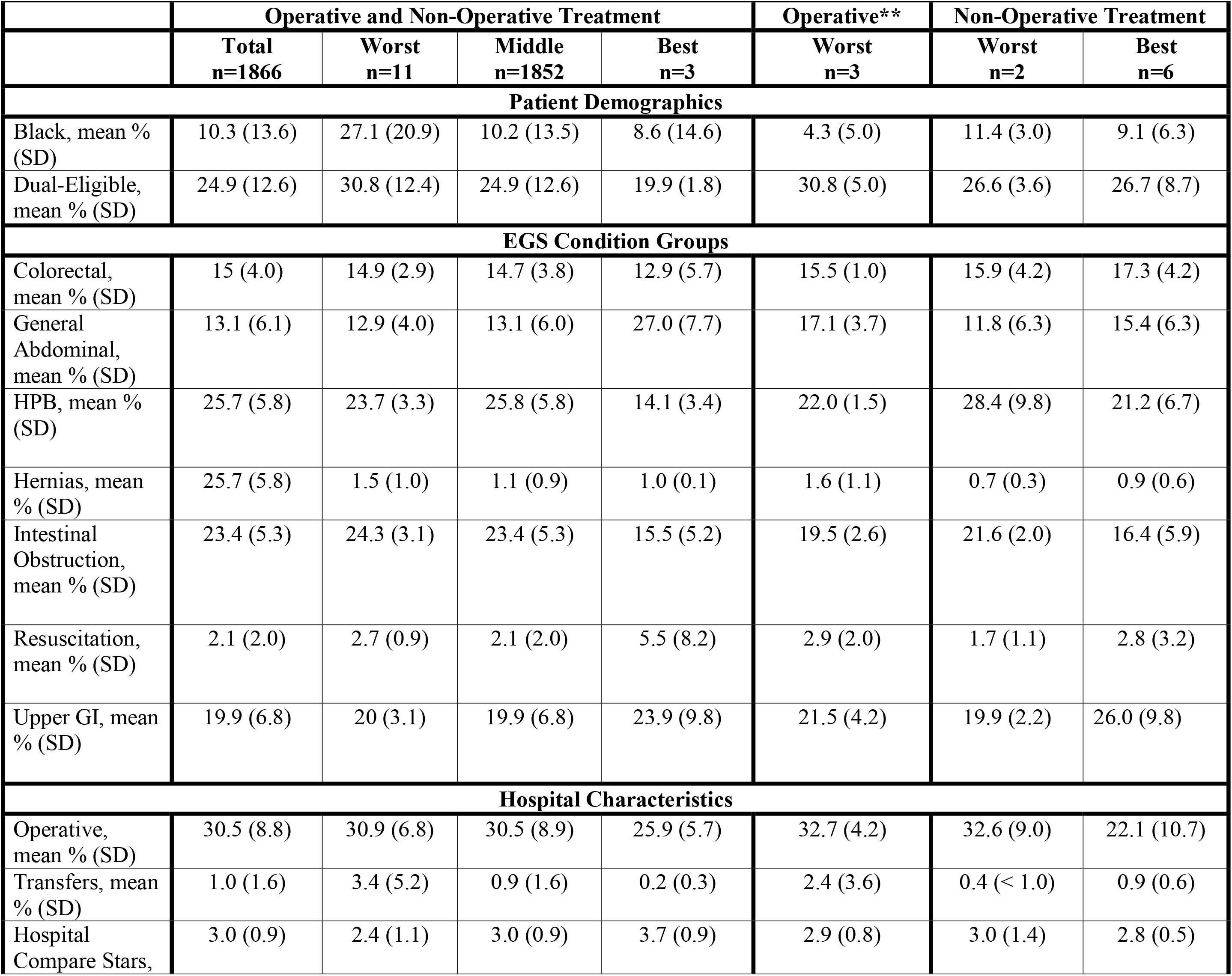
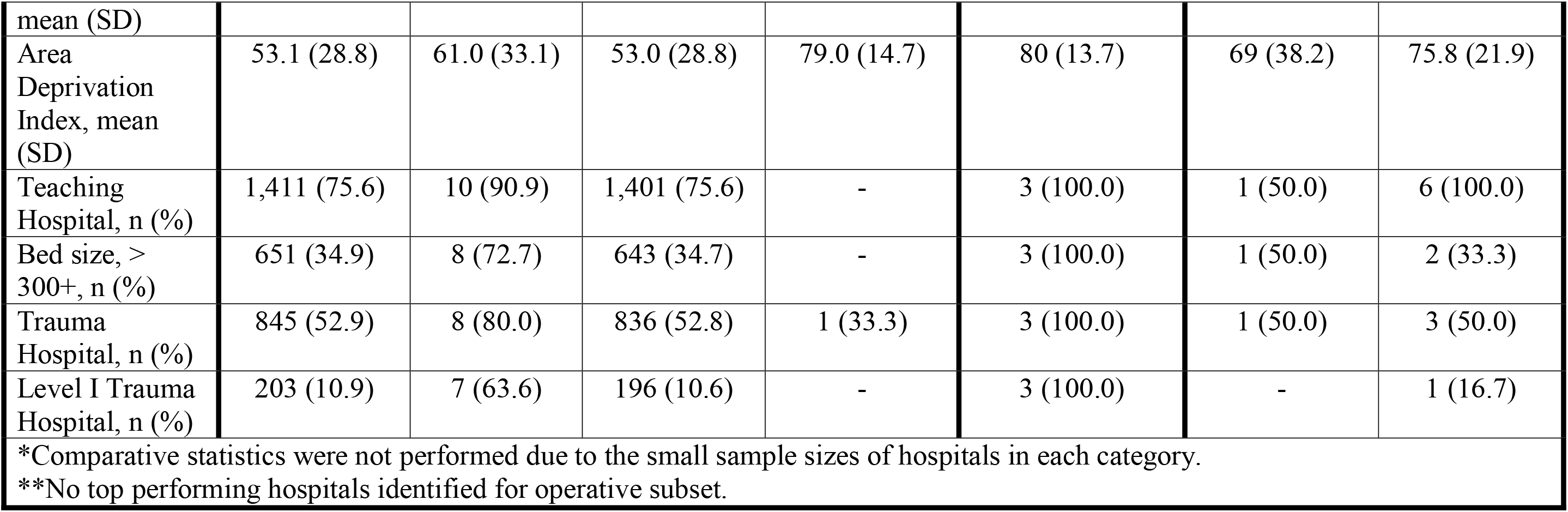
Patient and Hospital Characteristics Across Hospitals, by Treatment Group^*^.

|  | Operative and Non-Operative Treatment |  |  |  | Operative** | Non-Operative Treatment |  |
| --- | --- | --- | --- | --- | --- | --- | --- |
|  | Total<br>n=1866 | Worst<br>n=11 | Middle<br>n=1852 | Best<br>n=3 | Worst<br>n=3 | Worst<br>n=2 | Best<br>n=6 |
| Patient Demographics |  |  |  |  |  |  |  |
| Black, mean % (SD) | 10.3 (13.6) | 27.1 (20.9) | 10.2 (13.5) | 8.6 (14.6) | 4.3 (5.0) | 11.4 (3.0) | 9.1 (6.3) |
| Dual-Eligible, mean % (SD) | 24.9 (12.6) | 30.8 (12.4) | 24.9 (12.6) | 19.9 (1.8) | 30.8 (5.0) | 26.6 (3.6) | 26.7 (8.7) |
| EGS Condition Groups |  |  |  |  |  |  |  |
| Colorectal, mean % (SD) | 15 (4.0) | 14.9 (2.9) | 14.7 (3.8) | 12.9 (5.7) | 15.5 (1.0) | 15.9 (4.2) | 17.3 (4.2) |
| General Abdominal, mean % (SD) | 13.1 (6.1) | 12.9 (4.0) | 13.1 (6.0) | 27.0 (7.7) | 17.1 (3.7) | 11.8 (6.3) | 15.4 (6.3) |
| HPB, mean % (SD) | 25.7 (5.8) | 23.7 (3.3) | 25.8 (5.8) | 14.1 (3.4) | 22.0 (1.5) | 28.4 (9.8) | 21.2 (6.7) |
| Hernias, mean % (SD) | 25.7 (5.8) | 1.5 (1.0) | 1.1 (0.9) | 1.0 (0.1) | 1.6 (1.1) | 0.7 (0.3) | 0.9 (0.6) |
| Intestinal Obstruction, mean % (SD) | 23.4 (5.3) | 24.3 (3.1) | 23.4 (5.3) | 15.5 (5.2) | 19.5 (2.6) | 21.6 (2.0) | 16.4 (5.9) |
| Resuscitation, mean % (SD) | 2.1 (2.0) | 2.7 (0.9) | 2.1 (2.0) | 5.5 (8.2) | 2.9 (2.0) | 1.7 (1.1) | 2.8 (3.2) |
| Upper GI, mean % (SD) | 19.9 (6.8) | 20 (3.1) | 19.9 (6.8) | 23.9 (9.8) | 21.5 (4.2) | 19.9 (2.2) | 26.0 (9.8) |
| Hospital Characteristics |  |  |  |  |  |  |  |
| Operative, mean % (SD) | 30.5 (8.8) | 30.9 (6.8) | 30.5 (8.9) | 25.9 (5.7) | 32.7 (4.2) | 32.6 (9.0) | 22.1 (10.7) |
| Transfers, mean % (SD) | 1.0 (1.6) | 3.4 (5.2) | 0.9 (1.6) | 0.2 (0.3) | 2.4 (3.6) | 0.4 (< 1.0) | 0.9 (0.6) |
| Hospital Compare Stars, | 3.0 (0.9) | 2.4 (1.1) | 3.0 (0.9) | 3.7 (0.9) | 2.9 (0.8) | 3.0 (1.4) | 2.8 (0.5) |

Unadjusted results are presented in Table 2. A caterpillar plot (Figure 1) demonstrates individual hospital performance as measured by risk standardized adverse event rates relative to the remainder of the hospitals. There are only a select few hospitals that either performed statistically better (at the right tail) or worse than (at the left tail) the remainder of the hospital dataset. After adjustment, there was a significantly higher rate of adverse events at the worst hospitals (57% ± 4%)) when compared with the best hospitals (25% ± 5%). There were no best performing hospitals for operative treatment. The worst hospitals for operative treatment displayed adjusted rates of adverse events that were similar to those seen for the hospitals performing worst in non-operative treatment (57% ± 10%). For non-operative treatment, there was a higher rate of adverse events at the worst hospitals (60% ± 2%) when compared with the best hospitals (29% ± 6%). Secondary outcomes demonstrated similar findings. (Table 2)

**Table 2:**
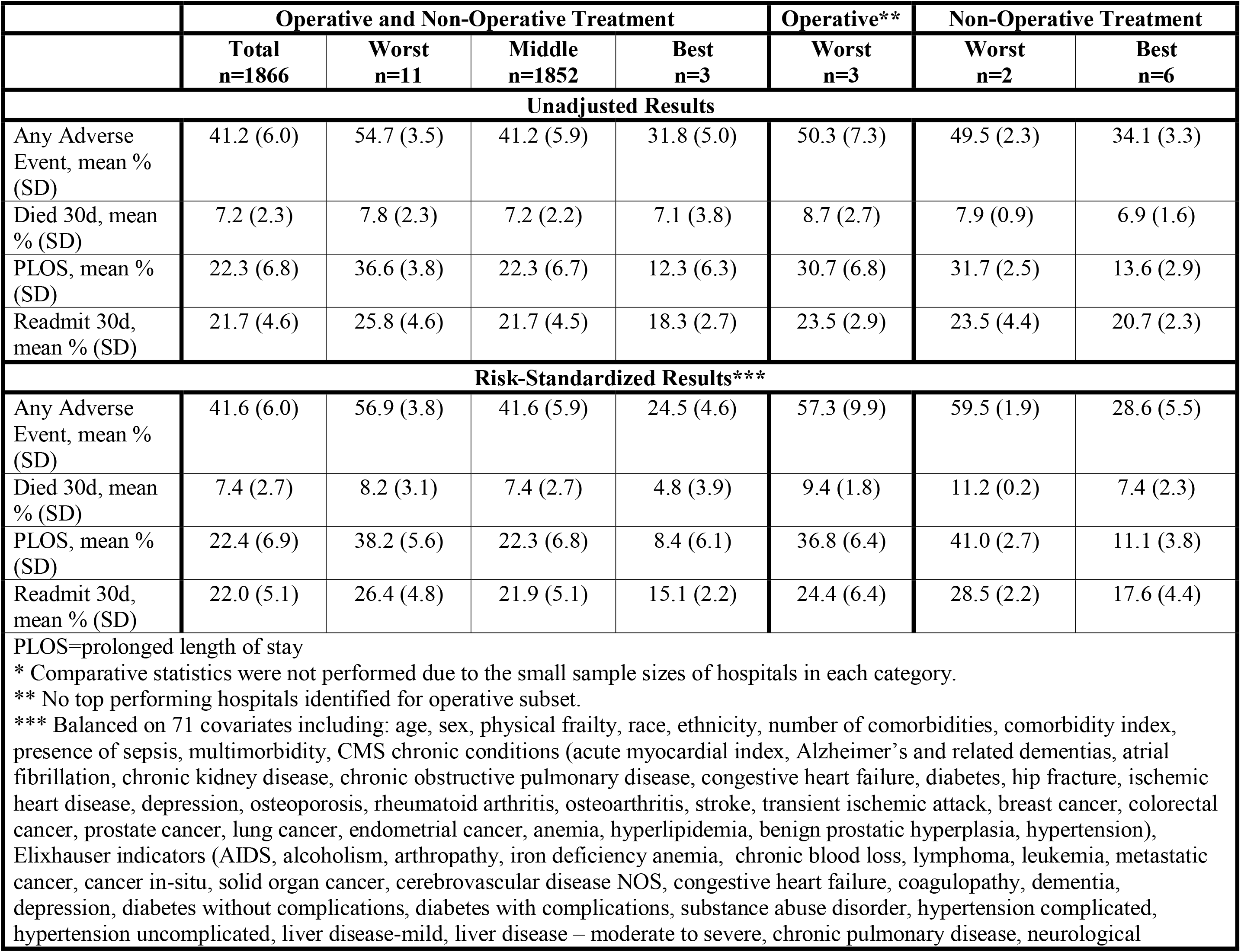
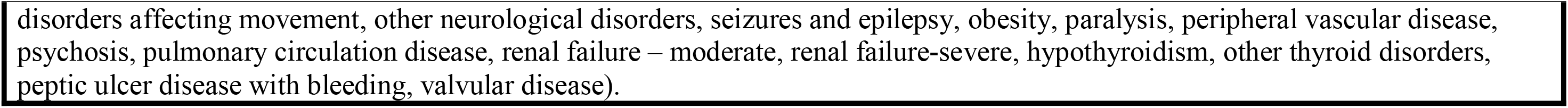
Outcomes by Hospital Ranking and Treatment Group*.

|  | Operative and Non-Operative Treatment |  |  |  | Operative** | Non-Operative Treatment |  |
| --- | --- | --- | --- | --- | --- | --- | --- |
|  | Total<br>n=1866 | Worst<br>n=11 | Middle<br>n=1852 | Best<br>n=3 | Worst<br>n=3 | Worst<br>n=2 | Best<br>n=6 |
| **Unadjusted Results** |  |  |  |  |  |  |  |
| Any Adverse Event, mean % (SD) | 41.2 (6.0) | 54.7 (3.5) | 41.2 (5.9) | 31.8 (5.0) | 50.3 (7.3) | 49.5 (2.3) | 34.1 (3.3) |
| Died 30d, mean % (SD) | 7.2 (2.3) | 7.8 (2.3) | 7.2 (2.2) | 7.1 (3.8) | 8.7 (2.7) | 7.9 (0.9) | 6.9 (1.6) |
| PLOS, mean % (SD) | 22.3 (6.8) | 36.6 (3.8) | 22.3 (6.7) | 12.3 (6.3) | 30.7 (6.8) | 31.7 (2.5) | 13.6 (2.9) |
| Readmit 30d, mean % (SD) | 21.7 (4.6) | 25.8 (4.6) | 21.7 (4.5) | 18.3 (2.7) | 23.5 (2.9) | 23.5 (4.4) | 20.7 (2.3) |
| **Risk-Standardized Results\*\*\*** |  |  |  |  |  |  |  |
| Any Adverse Event, mean % (SD) | 41.6 (6.0) | 56.9 (3.8) | 41.6 (5.9) | 24.5 (4.6) | 57.3 (9.9) | 59.5 (1.9) | 28.6 (5.5) |
| Died 30d, mean % (SD) | 7.4 (2.7) | 8.2 (3.1) | 7.4 (2.7) | 4.8 (3.9) | 9.4 (1.8) | 11.2 (0.2) | 7.4 (2.3) |
| PLOS, mean % (SD) | 22.4 (6.9) | 38.2 (5.6) | 22.3 (6.8) | 8.4 (6.1) | 36.8 (6.4) | 41.0 (2.7) | 11.1 (3.8) |
| Readmit 30d, mean % (SD) | 22.0 (5.1) | 26.4 (4.8) | 21.9 (5.1) | 15.1 (2.2) | 24.4 (6.4) | 28.5 (2.2) | 17.6 (4.4) |

PLOS=prolonged length of stay \* Comparative statistics were not performed due to the small sample sizes of hospitals in each category. \*\* No top performing hospitals identified for operative subset. \*\*\* Balanced on 71 covariates including: age, sex, physical frailty, race, ethnicity, number of comorbidities, comorbidity index, presence of sepsis, multimorbidity, CMS chronic conditions (acute myocardial index, Alzheimer’s and related dementias, atrial fibrillation, chronic kidney disease, chronic obstructive pulmonary disease, congestive heart failure, diabetes, hip fracture, ischemic heart disease, depression, osteoporosis, rheumatoid arthritis, osteoarthritis, stroke, transient ischemic attack, breast cancer, colorectal cancer, prostate cancer, lung cancer, endometrial cancer, anemia, hyperlipidemia, benign prostatic hyperplasia, hypertension), Elixhauser indicators (AIDS, alcoholism, arthropathy, iron deficiency anemia, chronic blood loss, lymphoma, leukemia, metastatic cancer, cancer in-situ, solid organ cancer, cerebrovascular disease NOS, congestive heart failure, coagulopathy, dementia, depression, diabetes without complications, diabetes with complications, substance abuse disorder, hypertension complicated, hypertension uncomplicated, liver disease-mild, liver disease – moderate to severe, chronic pulmonary disease, neurological
disorders affecting movement, other neurological disorders, seizures and epilepsy, obesity, paralysis, peripheral vascular disease, psychosis, pulmonary circulation disease, renal failure – moderate, renal failure-severe, hypothyroidism, other thyroid disorders, peptic ulcer disease with bleeding, valvular disease).

**Table 4:** Standard and Rank Correlations Between Operative and Non-Operative Performance Measures.

|  | **Standard Correlations** | **p-value** | **Spearman Rank Correlations** | **p-value** |
| --- | --- | --- | --- | --- |
| **30d Mortality** | 0.10 | <0.001 | 0.10 | <0.001 |
| **PLOS** | 0.27 | <0.001 | 0.32 | <0.001 |
| **Readmission** | 0.17 | <0.001 | 0.18 | <0.001 |
| **Adverse Event** | 0.20 | <0.001 | 0.21 | <0.001 |
| PLOS= prolonged length of stay |  |  |  |  |

**Figure 1:**
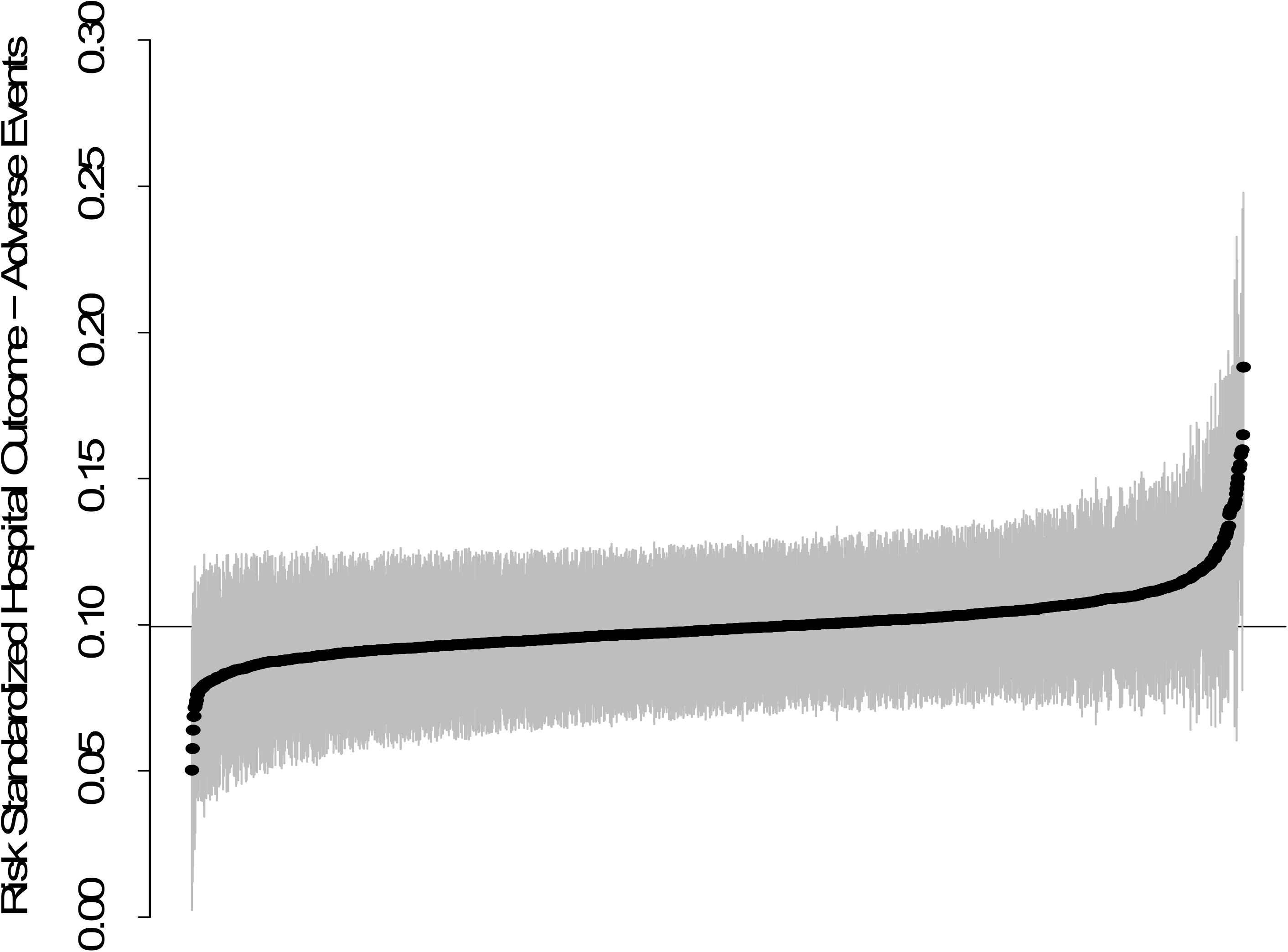
Caterpillar Plot Indicating Individual Hospital Risk-Standardized Adverse Event Rates.

The standard correlation between operative and non-operative outcomes at the hospital level was 0.10 for overall mortality, 0.27 for prolonged length of stay, 0.17 for readmission and 0.20 for adverse event. The Spearman rank correlation between the two subsets of patients was 0.10 for overall mortality, 0.32 for prolonged length of stay, 0.18 for readmission and 0.21 for adverse events (all p<0.001).

## Discussion

Using direct standardization to rank hospitals on EGS performance in this nationwide study of older adults, we demonstrate a more than two-fold difference in adverse event rates between the best and worst performing hospitals. Hospital performance differed similarly on 30-day death, prolonged length of stay and readmissions. There was also significant variation in hospital performance in the non-operative treatment subgroup. The worst hospitals for operative treatment achieved outlier status while there were no hospitals that distinguished themselves as the best in operative treatment.

Ingraham et al. found that EGS performance across conditions was only weakly correlated within hospitals. ^11^ Further, Zogg et al. demonstrated that within EGS conditions, hospital performance was consistent across multiple outcome types.^3^ We build on these findings to demonstrate that hospital performance on several metrics in operative and nonoperative treatment is only weakly correlated, and that there are a limited number of hospitals within the United States that are distinguished in EGS care of older adults.

## Limitations

This study has several limitations. First, variation in coding practices across hospitals may impact the capture of adverse events. However, for this study, we captured death, prolonged length of stay, and readmissions which are all known to be valid and reliable in Medicare claims. Second, direct standardization can only adjust for observable covariates which may explain the paucity of outlier hospitals. That is, if there are key patient covariates that are unobserved which contribute to hospital quality, we cannot account for these covariates under direct standardization. However, the number of best and worst performing hospitals in our study is similar to other published reports.^11,12^

## Conclusion

We have confirmed variation in EGS performance among hospitals treating older adults with the best hospitals significantly outperforming the worst hospitals on all outcome measures examined. These data suggest that EGS outcomes may be optimized with targeted hospital selection. Weak correlation between hospital performance on operative and non-operative care suggests that finding the right hospital for each patient is more complicated than defining a Center of Excellence designation. Further, over 14 years, there has only been a 13% increase in adoption of acute care surgery models that could improve EGS outcomes within hospitals.^13^ As such, alternative approaches to supplement quality and safety initiatives are needed until best practices become universal.

## Data Availability

All data produced in the present study are available upon reasonable request to the authors

## Acknowledgements

Research reported in this publication was supported by the National Institute on Aging of the National Institutes of Health under Award Number R01AG060612. The content is solely the responsibility of the authors and does not necessarily represent the official views of the National Institutes of Health.

## Notes

### Competing Interest Statement

The authors have declared no competing interest.

